# Central obesity, smoking habit and hypertension are associated with a blunted serological response to COVID-19 mRNA vaccine

**DOI:** 10.1101/2021.04.13.21255402

**Authors:** Mikiko Watanabe, Angela Balena, Dario Tuccinardi, Rossella Tozzi, Renata Risi, Davide Masi, Alessandra Caputi, Rebecca Rossetti, Maria Elena Spoltore, Valeria Filippi, Elena Gangitano, Silvia Manfrini, Stefania Mariani, Carla Lubrano, Andrea Lenzi, Claudio Mastroianni, Lucio Gnessi

**Affiliations:** Department of Experimental Medicine, Section of Medical Pathophysiology, Food Science and Endocrinology, Sapienza University of Rome, 00161 Rome, Italy; Department of Endocrinology and Diabetes, University Campus Bio-Medico of Rome, 00128 Rome, Italy; Department of Molecular Medicine, Sapienza University of Rome, 00161 Rome, Italy; Department of Public Health and Infectious Diseases, Sapienza University of Rome, 00185 Roma, Italy

**Keywords:** BMI, SARS CoV-2, infection, antibodies, waist circumference, vaccination

## Abstract

**Aims:** To explore variables associated with the serological response following COVID-19 mRNA vaccine.

**Methods:** Healthcare workers adhering to the vaccination campaign against COVID-19 were enrolled in January-February 2021. All subjects underwent two COVID-19 mRNA vaccine inoculations (Pfizer/BioNTech) separated by three weeks. Blood samples were collected before the first and 1-4 weeks after the second inoculation. Clinical history, demographics, and vaccine side effects were recorded. Baseline anthropometric parameters were measured, and body composition was performed through dual-energy-X-ray absorptiometry.

**Results:** Eighty-six patients were enrolled. Those with central obesity had lower antibody (Ab) titers compared with those with no central obesity [1426(1436)vs1971(1819), p=0.04]; smokers had a blunted response compared to non-smokers [1099(1350)vs1921(1375), p=0.007], as well as hypertensive vs normotensive [650±1192vs1911(1364), p=0.001] and dyslipidemic compared to those with normal serum lipids [534(972)vs 1872(1406), p=0.005]. Multivariate analysis showed that higher waist circumference, smoking, hypertension and longer time elapsed since second vaccine inoculation were associated with lower Ab titers, independent of BMI, age and gender. The association between waist circumference and Ab titers was lost when controlling for body fat, suggesting that visceral accumulation may explain this result.

**Conclusions:** It is currently impossible to determine whether lower SARS CoV-2 Abs lead to higher likelihood of developing COVID-19. However, neutralizing abs correlate with protection against several viruses including SARS-CoV-2, and the finding that central obesity, hypertension and smoking are associated with a blunted response warrants further attention. Our findings must lead to a vigilant approach, as these subjects could benefit from earlier vaccine boosters or different vaccine schedules.

## Introduction

Since Severe acute respiratory syndrome coronavirus 2 (SARS-CoV-2) was described in 2019, the world has faced an unprecedented pandemic causing millions of deaths. Obesity and excess visceral fat were shown to be major risk factors for the development of complications following COVID-19 infection ^1-4^, and, even before vaccines were made available, concerns regarding the possibility that obesity may blunt their efficacy were raised ^5^. In December 2021, Polack&Thomas et al. reported promising results regarding an mRNA vaccine against COVID-19, BNT162b2 (Pfizer, Inc and BioNTech), which conferred 95% protection against COVID-19 in adult subjects. The trial enrolled approximately 44 000 subjects, and there were 8 and 162 COVID-19 cases, respectively, following the two doses of vaccine or placebo. Despite the population comprising 35% of patients with a BMI>30, and the good efficacy data reported towards COVID-19 infection, evidence regarding the ability to protect against severe COVID-19 is less certain for subpopulations, and long-term data is lacking^6^. Moreover, obesity is not defined by BMI, but by the presence of fat excess^7^, BMI often mistakenly including subjects with fat excess in the normal weight category and *viceversa*. Body fat measured through Bioimpedentiometry (BIA) or Dual X-Ray Absorptiometry (DXA) may represent an alternative, but cut off values have never been validated, and the necessary equipment may not be promptly available in many clinical practices. The use of waist circumference as a measure of central obesity is, conversely, well established, and the European Association for the Study of Obesity (EASO), recommends to screen for obesity complications not only those with a BMI≥25, but also those with central obesity (waist circumference ≥80 cm for women and ≥ 94 cm for men). To the best of our knowledge, no studies are available to date investigating the real-life immunogenicity following BNT162b2 vaccine in relation to body composition and body fat distribution. In Italy, the BNT162b2 Pfizer BioNtech vaccine has been selected to be administered to healthcare professionals since December 2020. We aimed to explore variables associated with the response to the COVID-19 vaccine in a cohort of health care workers, focusing on adiposity parameters such as central obesity.

## Materials and methods

### Population and study design

Subjects included in this single center observational study were enrolled among health care workers of Policlinico Umberto I of Rome voluntarily undergoing a Pfizer BioNtech COVID-19 vaccine in January and February 2021. All subjects had undergone repeat naso- and oro-pharyngeal swabs as per hospital policy throughout the pandemic, and no previous infection had been recorded. Before enrollment, all subjects underwent a venous blood draw in order to confirm absence of antibodies against Sars-CoV-2. Those who had a positive serology were excluded from the study. The inclusion criteria were as follows: age over 18 years old, stable body weight (less than 5 kg self-reported change during the preceding 3 months); absence of previous SARS CoV-2 infection, absence of contraindications to the vaccine, willingness to undergo voluntary vaccination, absence of immunodepression, no use of medications known to impact the immune system and no ongoing pregnancy. Data about demographic characteristics were collected with the means of a structured interview. The study was approved by the local IRB (prot. CE 6228, Sapienza University of Rome), conducted in accordance with the Declaration of Helsinki and the Good Clinical Practice. Written informed consent was obtained from all study participants before enrollment.

### Vaccination procedure and blood collection

All patients were subjected to two COVID-19 vaccine inoculations, separated by 21 days (Comirnaty, Pfizer-BioNTech, Berlin, Germany). Before the first inoculation, all patients underwent a blood draw that was handled according to local standards of practice. A second blood draw was collected between one and four weeks after the second inoculation, 28 to 49 days after the first inoculation. Samples were centrifuged and plasma kept at -80°C until further analysis.

### Biochemical measures

Routine biochemical tests were handled according to standard operating procedures. Anti SARS Cov2 antibodies were measured through a commercially available assay (Elecsys® Anti-SARS-CoV-2 assay, Roche Diagnostics, Rotkreuz, Switzerland), which detects total antibodies against the SARS-CoV-2 spike (S) antigen in a sandwich electrochemiluminescence assay (ECLIA) ^8^.

### Anthropometric and body composition assessment

Anthropometric parameters were measured at baseline. Body weight was measured using a balance-beam scale (Seca GmbH & Co, Hamburg, Germany). Height was rounded to the closest 0.5 cm. BMI was calculated as weight in kilograms divided by squared height in meters (kg/m^2^). Waist circumference was measured midway between the lower rib and the iliac crest, hip circumference at the level of the widest circumference over the great trochanters to the closest 1.0 cm. The measurements were performed with the means of an anelastic tape by trained professionals. Body composition was measured through dual-energy-X-ray absorptiometry (DXA) (Hologic 4500, Bedford, MA, USA) as previously reported ^9^.

### Statistics

The Statistical Package for Social Sciences (SPSS), v.20 was used for statistical analysis. Results are presented as mean, standard deviation (SD) or median, Interquartile Range (IQR) according to their distribution. Normality was assessed with the Kolmogorov–Smirnov test. Variables not normally distributed were log-transformed. A Kruskal-Wallis test with a Bonferroni post-hoc multiple comparison was conducted to compare the distribution of the COVID-19 Ab titers among the four different levels (quartiles) of waist circumference. A Mann-Whitney U test was conducted to compare the distribution of the COVID-19 Abs in subpopulations. Univariate and multivariate linear regression models were performed to analyze the relationship between the SASR CoV-2 Ab as the dependent variable and clinical, biochemical, DEXA-derived body composition parameters as the independent variables.

To build a multivariate linear regression model with Ab titers as the dependent variable, we used an enter method approach (all the independent variables included in the same regression equation) and investigated the following variables/models: (1) multivariate analysis including age and BMI together with variables with significant univariate association (p value ≤.05) analyzed one by one as regressors (Age + BMI + Waist Circumference; Age + BMI + Waist to Hip Ratio; Age + BMI + Central Obesity; Age + BMI + Time; Age + BMI + Hypertension; Age + BMI + Dyslipidemia; Age + BMI + Smoking habit). (2) Multivariate analysis including age, BMI and Time since second inoculation together with variables with significant association (p value ≤.05) at multivariate model 1, analyzed one by one as regressors (Age + BMI + Time since second inoculation + Waist Circumference; Age + BMI + Time since second inoculation + Hypertension; Age + BMI + Time since second inoculation + Dyslipidemia; Age + BMI + Time since second inoculation + Smoking habit). (3) Multivariate analysis including all statistically significant variables of the multivariate model 1 with the addition of gender, age and BMI, as regressors in one single model. (4) Multivariate analysis including all statistically significant variables of the multivariate model 1 with the addition of gender, age, BMI, and total body fat mass (quartiles) as regressors in one single model. The variables results were added in the table, reporting their B and 95% CI, [R^2^]. For the analysis, a P-IN=0.05 and a P-OUT=0.10 were used. The effect estimate is reported as the coefficient of determination R^2^, which informs on how much the model explains the variance of the dependent variable. Variance inflation factor (VIF) values were lower than 4.0, suggesting the absence of multicollinearity between included variables ^10^. The results were considered statistically significant when p < 0.05.

## Results

### Study Population

Eighty-six subjects were enrolled in the present study in January and February 2021 among health care workers adhering to the vaccination campaign of Policlinico Umberto I Hospital, Rome, Italy. The clinical characteristics of the participants are summarized in **Table 1**. Briefly, the age was 29 (17), 39.5% male, BMI 22.4 (5.5) kg/m^2^, all were caucasian. 31.7% was a current smoker, 15.3% was hypertensive on pharmacological treatment (of which almost 100% on angiotensin-converting enzyme inhibitors/ angiotensin II receptor blockers, ACEI/ARBs), only 2.4% was diabetic and 7.1% dyslipidemic. 76.8% had undergone routine influenza virus vaccination in the preceding 12 months, as recommended per hospital policy. A small panel of routine biochemical tests was normal for all participants and is summarized in **Table 1**. Regarding adiposity measures, 63.1% had a normal weight, 27.4% overweight and 9.5% obesity according to BMI cut-offs. However, central obesity was observed in 60.9%(n=53) of subjects out of 78 for whom waist circumference measurements had been recorded. The cutoff to determine central obesity was 80 cm for women, and 94 cm for men^11^.

**Table 1.** Descriptive characteristics of study population

|  | Mean±SD or Median(IQR) or N(%) |
| --- | --- |
| Age (years) | 29(17) |
| Weight (kg) | 66(17.8) |
| Gender (%F) | 60.50% |
| BMI (kg/m<sup>2</sup>) | 22.4(5.5) |
| Waist circumference (cm) | 91±11.3 |
| Hip Circumference (cm) | 99±9 |
| Waist-to-hip ratio | .9(.09) |
| Fat mass (kg) | 16.1(7.6) |
| Lean mass (kg) | 48.4(16.3) |
| Time (days)* | 22(5) |
| Smoking habit N(%) | 26(31.7%) |
| Hypertension N(%) | 13(15.3%) |
| Type 2 Diabetes N(%) | 2(2.4%) |
| Dyslipidemia N(%) | 6(7.1%) |
| Creatinine (mg/dL) | .86(.25) |
| BUN (mg/dL) | 30.6±8.7 |
| ALT(U/L) | 15(13.5) |
| AST(U/L) | 21(9) |
| Total cholesterol (mg/dL) | 200(55) |
| HDL cholesterol (mg/dL) | 58(26) |
| Triglycerides (mg/dL) | 85(71) |
| Uric acid (mg/dL) | 4.5(1.8) |
| CRP (mg/L) | 700(1100) |

Notes: IQR, Interquartile range; F, female; N, number; BUN, Blood urea nitrogen; ALT, Alanine aminotransferase; AST, Aspartate aminotransferase; CRP, C-reactive protein.

### Safety

All adverse events following the vaccine inoculation were recorded with the means of a structured interview, and 65.9% (n=56) of participants complained of some adverse events following the first inoculation. Of these, 50 reported of pain or pruritus in the site of inoculation, 10 of headache, fatigue or malaise, 3 of low-grade fever, 2 of dyspnea and 5 of other minor adverse events. Following the second inoculation, 78.2% (n=61) reported some adverse event, of which 44 reported of pain or pruritus in the site of inoculation, 28 of headache, fatigue or malaise, 21 of low-grade fever, and 8 of other minor adverse events. No major adverse event requiring hospitalization was recorded at any time point. Adiposity parameters such as higher waist circumference, waist-to-hip ratio, BMI, central obesity or body fat were not associated with more adverse events (data not shown).

### Efficacy

Patients with central obesity had significantly lower SARS CoV-2 antibody titers compared with those with no central obesity [1426 (1436) vs 1971 (1819), respectively, p=0.04; **Figure 1A**] Similarly, there was a significant difference regarding Ab titers depending on the waist circumference quartile subjects belonged to (p=0.046). Post hoc analysis showed that those belonging to the second quartile had significantly higher Ab titers compared to those belonging to the fourth waist circumference quartile [p=0.036; **Figure 1 B**]. Interestingly, obesity identified as a BMI≥ 30 Kg/m^2^ was not associated with a blunted response [p=0.524; data not shown]. Furthermore, subjects with a smoking habit had a blunted response compared to those who were not current smokers [1099 (1350) vs 1921 (1375), respectively, p=0.007; **Figure 1 C**], and the same was for those who were hypertensive compared to those who were not [650±1192 vs 1911(1364), respectively, p=0.001; **Figure 1 D**], and those with dyslipidemia compared to those who had a normal lipid profile and were not on lipid lowering drugs [534 (972) vs 1872 (1406), respectively, p=0.005; **Figure 1 E**].

**Figure 1.**
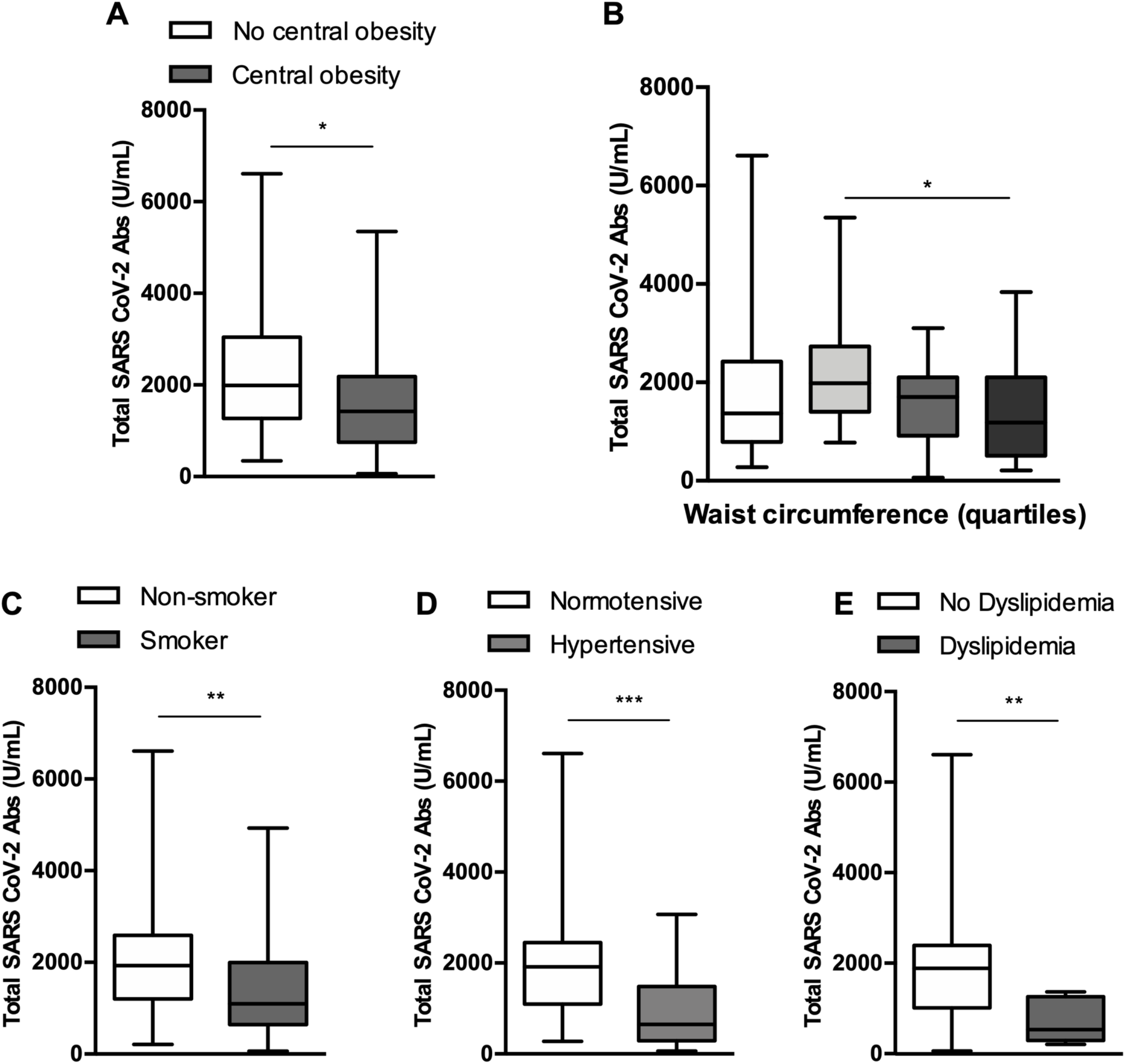
Serological response of analyzed subpopulations. Anti SARS CoV-2 Antibody titers were significantly lower in subjects with central obesity **(A)**, subjects belonging to the second quartile of waist circumference compared to those belonging to the fourth quartile (overall p= .046) **(B)**, smokers **(C)**, hypertensive subjects **(D)**, dyslipidemic subjects **(E)**. *p<.05, **p<.01, **p<.001, ***p<.000

Regression analysis showed that the presence of central obesity (as dichotomic variable, waist-to-hip ratio or crude waist circumference measurement) was associated with a blunted serological response to the vaccine, as were hypertension, dyslipidemia, and smoking habit **(Table 2)**. Moreover, the time since the second vaccine inoculation at which the Ab titers were evaluated was significantly associated with a decline in serum SARS CoV-2 Ab **(Table 2)**. The presence of side effects following the first or second inoculation was not associated to different Ab titers, neither was flu vaccination in the preceding 12 months **(Table 2)**.

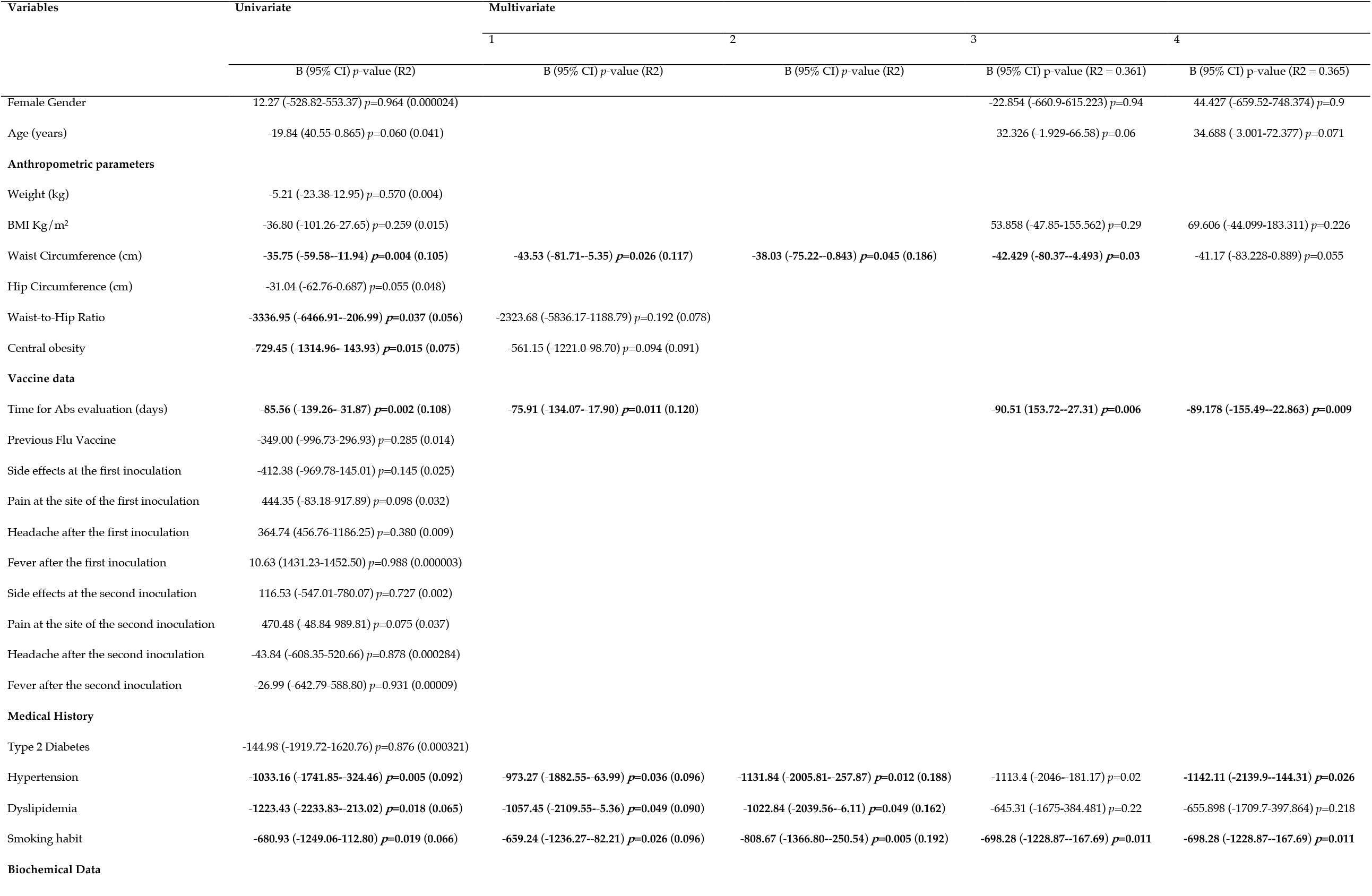
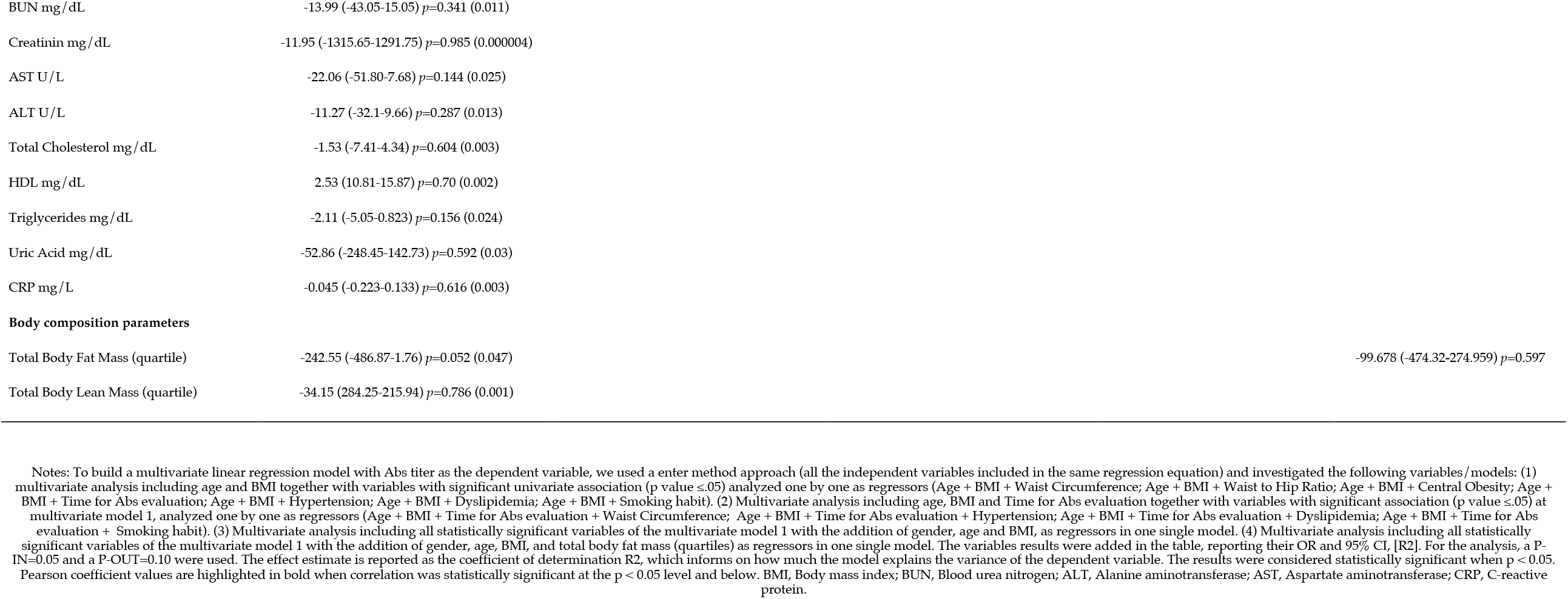

Each parameter showing a significant univariate association with Ab titers were included in a multivariate analysis together with age and BMI, factors possibly affecting the association, analyzed one by one as regressors **(Table 2, multivariate model 1)**. Waist circumference, time elapsed since vaccination, hypertension, diabetes and dyslipidemia retained a significant association. As the time factor was strongly associated with the Ab titers, we conducted a second multivariate analysis including each parameter with a significant association at multivariate model 1, together with BMI, age and time as factors likely playing a relevant role in the association, analyzed one by one as regressors **(Table 2, multivariate model 2)**. The addition of time in the model did not influence the results. A third multivariate model including all the statistically significant variables of the multivariate model 1 together, controlling for gender, age and BMI, showed that waist circumference, time since vaccination, hypertension and smoking habit were still significantly associated with Ab titers **(Table 2, multivariate model 3)**. To test whether body fat underlay the significant association between waist circumference and the blunted serological response to vaccination, a fourth multivariate model included DXA measured body fat on top of all parameters included in model 3, showing that indeed the association of waist circumference was lost. Conversely, the association with hypertension and smoking habit were maintained **(Table 2, multivariate model 4)**.

## Discussion

Herein, we report that central obesity, independent of BMI, but dependent of body fat mass, is associated with lower Ab titers following a COVID-19 mRNA vaccine. This could be due to a number of reasons, one of which is the metabolic derangements that often come with visceral adiposity, together with the immune dysfunction that has been reported in patients with obesity^12,13^. In fact, strong evidence supports the fact that obesity is also associated with poor seroconversion upon some vaccine administration^14^, together with increased risk of infection even when the seroconversion seems robust ^15^. A recent study has shown that higher BMIs are associated with lower serological responses after COVID-19 vaccine in Italian healthcare workers^16^. Although pointing in the same direction, our findings slightly differ from this study, as we could not find any significant association between BMI and SARS CoV-2 ab titers following vaccination. This could be due to a narrower distribution in terms of BMI in our cohort, but it should also be highlighted that the kits to detect Ab titers were different, as was the timing of the Ab measurement (one week). Noteworthy, Ab titers tend to rapidly decline following the vaccine with a kinetics not entirely elucidated yet and potentially different in some subpopulations^17^. Moreover, it is acknowledged that BMI poorly describes actual body fat excess, which is the definition of obesity according to the World Health Organization^7^, and a simple parameter such as waist circumference is shown to be even more associated to chronic low grade inflammation and cardiovascular disease, morbidity and mortality as opposed to BMI^18^. Moreover, we have recently shown that visceral fat is the strongest predictor of the need of intubation following COVID-19 infection^1^, suggesting that the same trend might apply to the vaccine response, where an accumulation in central fat may impact Ab levels, possibly hindering the response of these patients against an eventual subsequent infection. It is therefore of utmost importance to determine in future studies whether cell mediated immunity is maintained even when Ab titers decline in this population.

We also report that hypertension as well as smoking were strongly associated with lower Ab titers. The present study was not specifically designed to investigate these aspects, and further studies are warranted to explore whether hypertension, the use of certain anti-hypertensive medications, and smoking are associated with poor response. Interestingly, it was previously reported that Ab titers following influenza vaccination decline more rapidly in smokers, through an unknown mechanism^19^. More generally, the habit of smoking is associated with a dysfunctional immune system, linked with both autoimmune disease and reduced response to infections^20^. Similarly, hypertension and an inappropriate response to vaccinations might have a common root into a dysfunctional immune system according to recent evidence ^21^. To date, no association between smoking or hypertension and reduced protection following COVID-19 vaccine has been reported.

Our study has several limitations. First, the time of evaluation ranged between 1 and 4 weeks following the second vaccine inoculation, introducing time as a possible bias. However, when the evaluations were controlled for this factor, they still yielded the same results. Second, the BMI range distribution was relatively narrow, although reflecting the prevalence of overweight and obesity in the general Italian population^22^. This could have hampered possible significant results regarding BMI differences. However, we collected other relevant adiposity parameters, such as DXA derived body fat and waist and hip circumference, which showed a wide distribution within our study population. Further, the sample size was relatively small, and patients with multiple comorbidities were underrepresented, possibly hindering some of the results. This was a study investigating the Ab titers shortly after the inoculation, and studies following the same patients over time are warranted in order to investigate the Ab kinetics according to body composition and other possibly relevant factors. Moreover, cell mediated immunogenicity warrants further attention, and studies including these outcomes are therefore needed. Certainly, anti SARS CoV-2 Ab titers following vaccination cannot predict the likelihood of developing COVID-19 at this point, and low but measurable levels may as well be highly protective against infection. However, neutralizing Ab titers correlate with protection against several viruses including SARS-CoV-2 ^23,24^, and the finding that central obesity, hypertension and smoking are associated with a blunted serological response shortly after the vaccination warrants further attention, as this may mean that these subjects respond in a different way to the same vaccination and may require different vaccine booster schedules over time.

Our study also features some strengths. This is, to the best of our knowledge, the first study reporting data on the immunogenicity of a COVID-19 vaccine according to central obesity indices. Healthcare professionals were the first being vaccinated across all countries, so these are the earliest real-life findings becoming available. Waist circumference as a marker of central obesity does not require additional instrumental tests, it is cheap and easy to collect, and it therefore possesses a possible immediate clinical applicability. Clinical history was acquired with the means of a standardized structured interview allowing for a thorough and complete collection, and the adverse events were reported three days after the two vaccine inoculations, limiting the risk of recall bias.

With the general population now being vaccinated, more and more subjects with central and general obesity will receive the vaccine, and very soon booster schedules will need to be planned. The fact that the Ab response is blunted in certain subjects shortly after the second inoculation must lead to a highly vigilant approach, as medium and long-term data will become available only when the schedule will have been necessarily set already.

## Data Availability

Alla data is available upon reasonable request to the corresponding author

## Contributors

MW, AB, and DT collaborated equally on this work and are joint first authors. MW, AB, DT, SiM, StM, CL, AL, CM, LG contributed to the conception and design of the work. MW coordinated the work, supported by SiM, StM, CL, AL, CM, LG. DT, RenR, CL conducted the statistical analysis. All authors provided substantial scientific input in interpreting the results, drafting and or reviewing the manuscript. MW is the guarantor. The corresponding author attests that all listed authors meet authorship criteria and that no others meeting the criteria have been omitted.

## Ethical approval

The study was approved by the local IRB (prot. CE 6228), conducted in accordance with the Declaration of Helsinki and the Good Clinical Practice. Written informed consent was obtained from all study participants before enrollment.

